# Education, Cognitive Reserve, and Brain Pathology in Aging and Alzheimer’s Disease: Evidence from Preclinical and Symptomatic Cohorts

**DOI:** 10.64898/2026.05.07.26352659

**Authors:** Shima Raeesi, Amelie Metz, Katherine Chadwick, Yashar Zeighami, The PREVENT-AD Research Group, The Consortium for the Early Identification of Alzheimer’s Disease - Quebec (CIMA-Q), Cassandra Morrison, Mahsa Dadar

## Abstract

**INTRODUCTION:** Educational attainment is a key proxy of cognitive reserve (CR), but whether its association with cognition persists after accounting for multimodal brain pathology remains unclear.

**METHODS:** Data from PREVENT-AD and CIMA-Q were analyzed using cross-sectional, longitudinal, and residual-based models adjusted for multimodal pathology, including MRI-based white matter hyperintensities and atrophy measures, and plasma Alzheimer’s disease biomarkers. Education was examined in both cohorts. A multidimensional CR questionnaire was also examined in CIMA-Q.

**RESULTS:** Higher education was associated with better performance across multiple cognitive domains after accounting for pathology (p < 0.05). The CR composite score showed less consistent associations, and non-education subdomains showed limited added value. Longitudinally, education was more consistently associated with baseline cognitive levels than cognitive change, with executive-function trajectory differences observed in PREVENT-AD.

**DISCUSSION:** Findings support a CR framework in which education relates to better cognition despite pathology, while effects on decline are limited and domain-specific.

## 1. Background

Cognitive aging is marked by substantial inter-individual variability. Some older adults maintain relatively preserved cognitive function despite advancing age and accumulating brain pathology, whereas others experience more rapid deterioration^1–5^. This discrepancy between pathological burden and clinical expression is central to the concept of cognitive reserve (CR), which refers to the capacity to sustain cognitive performance despite age- or disease-related brain changes^3,6–10^. Within this framework, resilience mechanisms refer to processes that reduce the clinical impact of pathology, allowing individuals with comparable levels of brain pathology to show different levels of cognitive performance^3,6,7^. CR is most commonly studied using proxy indicators. Educational attainment is the most widely used proxy, reflecting early-life intellectual enrichment, and is reliably linked to better cognitive performance in aging and Alzheimer’s disease (AD) populations^8,11–21^. However, education captures only one dimension of CR^6,22^. Multidimensional CR questionnaires have therefore been developed to capture broader lifetime experiences, including occupational attainment, cognitively stimulating activities, language use, and social or leisure engagement^11,23,24^.

Prior studies suggest that CR may shape the relationship between brain pathology and cognition^12–15,17,21,25^. Higher education has been associated with weaker associations between AD-related hypometabolism and memory decline, as well as more efficient functional network connectivity^15,21^, suggesting that reserve may support cognition by improving the efficiency or flexibility of neural networks. Similarly, education may attenuate the cognitive impact of medial temporal lobe atrophy on episodic memory, although these effects appear to vary across disease stage, pathology severity, and cognitive domain^12,14,16,26^. Multidomain CR measures have also been associated with differences in how gray matter atrophy and white matter damage relate to cognition, supporting the idea that different components of reserve may contribute to resilience in distinct ways^7,11,25^. However, findings across studies remain heterogeneous, with evidence for both attenuating effects (i.e., reduced cognitive impact of pathology) and more complex, stage-dependent patterns of CR^3,11,14,16,27^. In particular, some studies suggest that individuals with higher CR may tolerate greater pathology before clinical symptoms emerge, although the impact on subsequent cognitive decline remains unclear^15,17,18^.

Despite substantial progress, important gaps remain. First, much of the literature examines CR in relation to single pathology markers^6,8,12,13^, although AD-related cognitive decline likely reflects the combined influence of vascular injury, neurodegeneration, and AD proteinopathies^3,8,28,29^. Studying these processes together is important as focusing on one marker at a time may obscure the cumulative or interacting contributions of multiple pathological pathways. Second, few studies examine CR across cohorts representing different stages of the AD continuum^14,21^. This limits understanding of whether reserve-related associations are similar in preclinical populations and in more clinically heterogeneous symptomatic groups. Third, it remains unclear whether CR-related associations are most evident for cognitive level, such as baseline performance, or whether they also extend to longitudinal trajectories of cognitive change after accounting for multimodal pathology^3,6,8,27^. This distinction is important because a reserve effect on cognitive level does not necessarily imply slower decline over time.

To address these gaps, the present study examined associations between CR proxies and cognitive performance in two aging cohorts representing complementary stages of the AD continuum. Specifically, this study tested whether education, as the primary CR proxy available in both cohorts, and a multidimensional CR questionnaire, available in one cohort, were associated with cognitive performance after accounting for cerebrovascular, neurodegenerative, and AD-related pathology markers. Residual-based analyses were also used to estimate cognition relative to pathology-predicted performance. It was hypothesized that higher CR would be associated with better cognitive performance despite measured brain pathology. By integrating cross-sectional, longitudinal, and residual-based approaches, this study further examined whether CR-related associations were more evident for baseline cognitive level or for trajectories of cognitive change over time.

## 2. Methods

### 2.1 Study populations

Data from two independent cohorts based in Quebec, Canada, representing complementary stages along the AD continuum were analyzed, enabling examination of CR across preclinical and symptomatic phases. Participants in both cohorts provided written informed consent, and study protocols were approved by the relevant institutional ethics committees.

The PREVENT-AD (PResymptomatic EValuation of Experimental or Novel Treatments for Alzheimer’s Disease)^30,31^ cohort is a single-site longitudinal study conducted at McGill University and designed to investigate individuals at increased risk for sporadic AD. All participants were required to have normal cognitive functioning at baseline, as determined by clinical evaluation and standardized neuropsychological testing, and were enriched for AD risk based on a family history of Alzheimer-like dementia in a first-degree relative^32,33^. Participants had to be at least 60 years old to be eligible or between 55 and 59 years old if they were within 15 years of the parental age at the onset of symptoms. Additional inclusion criteria included adequate visual and auditory capacities to complete testing and fluency in English or French. Exclusion criteria were the presence of significant neurological disorders (other than preclinical AD), major psychiatric illness, uncontrolled systemic medical conditions, or use of medications known to significantly affect cognition. Individuals with Magnetic Resonance Imaging (MRI) contraindications were also excluded. The PREVENT-AD protocol consists of annual follow-up visits with multimodal assessments, including detailed neuropsychological testing, MRI, and collection of blood and cerebrospinal fluid (CSF) biomarkers.

The CIMA-Q (Consortium for the Early Identification of Alzheimer’s Disease-Quebec)^34^ cohort is a multicenter longitudinal study that includes a wide clinical spectrum of aging and AD across multiple sites in Quebec. Participants with normal cognition, subjective cognitive decline (SCD), mild cognitive impairment (MCI), and mild AD dementia were recruited through memory clinics, community ads, and pre-existing research cohorts. Diagnostic classification was established using standardized clinical criteria, such as National Institute on Aging-Alzheimer’s Association (NIA-AA) guidelines^35,36^, based on clinical evaluation, neuropsychological testing, and, when available, biomarker information. Eligibility for the CIMA-Q study required participants to be aged 65 years or older, able to complete clinical and cognitive assessments, proficient in French or English, and have no contraindications to MRI. Individuals with severe systemic illnesses that could impair cognitive function, major psychiatric disorders, or significant neurological conditions unrelated to AD (e.g., stroke with major residual deficits, Parkinson’s disease) were excluded^34,37^. The CIMA-Q cohort also includes extensive phenotyping with clinical, neuropsychological, neuroimaging, and biomarker data^34^. Compared to PREVENT-AD, which focuses on preclinical individuals, CIMA-Q includes a more heterogeneous population with diverse degrees of cognitive impairment and increased pathological burden.

### 2.2 Measures

#### 2.2.1 Cognitive reserve (CR)

In this study, CR was assessed using three complementary approaches: (1) years of education as the primary proxy measure of CR, consistent with prior studies^8,12–20,26^; (2) a multidimensional CR questionnaire; and (3) a residual-based measure reflecting better-than-expected cognitive performance relative to pathology burden (defined later). Education, measured as years of formal schooling, was modeled as a continuous variable across all analyses in both cohorts. In the CIMA-Q cohort, CR was additionally assessed using the Bartrés Cognitive Reserve Questionnaire^38^, a validated multidomain instrument designed to capture lifetime cognitive enrichment across key intellectual and lifestyle domains. The questionnaire comprises eight items assessing formal education, parental education, participation in training courses, occupational attainment, musical training, language proficiency, reading habits, and engagement in cognitively stimulating activities (e.g., puzzles, games), reflecting factors associated with the development of cognitive reserve across the lifespan. Each item is scored categorically, and the total score (range 0-25) reflects cumulative lifetime CR, with higher scores indicating greater CR. To further characterize the multidimensional structure of CR, exploratory analyses were conducted by grouping questionnaire items into domain-specific composites reflecting cognitive enrichment-related experiences (musical engagement, training courses, and intellectual activities), social engagement (social participation and volunteer-related activities) and language proficiency (multilingual ability). These domain-level measures were derived by averaging scores of relevant items and were used to examine domain-specific contributions to cognitive performance.

#### 2.2.2 Cognition

Cognitive outcomes varied across cohorts depending on measures available. These measures were selected to ensure coverage of key cognitive domains relevant to aging and AD and maximize consistency and comparability between the two cohorts despite differences in available instruments. Global cognition was assessed using the Montreal Cognitive Assessment (MoCA)^39^ at baseline for both cohorts and for longitudinal assessment in the CIMA-Q cohort. MoCA is a widely used screening tool that evaluates multiple cognitive domains, including memory, executive function, attention, language, and visuospatial abilities. Longitudinal global cognitive performance in PREVENT-AD was evaluated using the Repeatable Battery for the Assessment of Neuropsychological Status (RBANS)^40^, which provides standardized measures of both global cognition and domain-specific performance, including immediate memory, delayed memory, attention, language, and visuospatial/constructional abilities. Additional domain-specific assessments included episodic memory, measured using the Rey Auditory Verbal Learning Test (RAVLT)^41,42^, which assesses immediate recall, delayed recall, and recognition memory across repeated trials. Given that several RBANS and RAVLT versions of potentially differing difficulty levels^43,44^ were used across visits, RBANS and RAVLT versions were included as covariates in the corresponding analyses to account for potential test-related variability. Executive function and processing speed were also assessed using the Trail Making Test (TMT)^45,46^. In most cognitive measures, higher scores indicate better performance; however, for TMT, higher scores reflect worse performance. Therefore, TMT scores were <u>reverse-coded</u> such that higher values consistently represent better cognitive performance across all outcomes.

#### 2.2.3 Brain pathology

Brain pathology was grouped into three complementary domains reflecting distinct but interacting processes relevant to AD: (1) cerebrovascular pathology, (2) neurodegeneration, and (3) AD-related pathology. This classification was used to capture the major biological pathways contributing to cognitive decline. Cerebrovascular pathology was indexed using total white matter hyperintensity (WMH) volumes derived from Fluid-Attenuated Inversion Recovery (FLAIR) MRI, reflecting small-vessel disease burden. Neurodegeneration was assessed using deformation-based morphometry (DBM; see MRI processing section for details), with a primary focus on medial temporal lobe structures, including the hippocampus, parahippocampal gyrus, and entorhinal cortex, as markers of AD-related neurodegeneration. Ventricular DBM measures were also included as indicators of more widespread global brain atrophy. AD-related pathology was evaluated using blood-based biomarkers, including plasma phosphorylated tau (p-tau231 and p-tau181, available in both CIMA-Q and PREVENT-AD), plasma Aβ42/Aβ40 ratio (available only in PREVENT-AD), and p-tau217 (available only in CIMA-Q). These biomarkers were selected based on their established sensitivity to amyloid and tau pathology and their relevance to early disease processes^47,48^. CSF biomarkers were not included due to limited availability and reduced sample size across cohorts, with CSF data available for approximately 60 participants in CIMA-Q and 140 in PREVENT-AD, which would have substantially reduced the sample size. All pathology measures were treated as continuous variables. To ensure consistent interpretation across markers, the medial temporal lobe DBM measures and Aβ42/Aβ40 ratio were <u>reverse-coded</u> such that higher values reflect greater pathological burden.

### 2.3 MRI processing

T1-weighted and FLAIR MRIs were processed using PELICAN^49^, a widely used^50–57^ open-source neuroimaging pipeline developed for analyzing large-scale multimodal datasets. Preprocessing involved denoising, intensity non-uniformity correction, and intensity normalization to a standardized range^58^.

Images were first linearly registered to a common stereotaxic space, followed by nonlinear registration to the MNI-ICBM152-2009c template. Jacobian determinants of the nonlinear deformation fields were used to derive DBM maps, providing information on local volumetric expansion or contraction at a voxel level. Voxel-wise DBM values were first averaged within anatomical regions using the CerebrA atlas^59^ to obtain regional DBM measures. These regional values were then log-transformed to improve normality and interpretability, such that negative values reflect local tissue loss (atrophy), whereas positive values indicate expansion (e.g., in the ventricles and sulci). Regional DBM measures were extracted for medial temporal lobe structures and ventricular regions, corresponding to markers of regional and global neurodegeneration described above. For consistency across imaging markers, DBM measures were <u>reverse-coded</u> such that higher values reflect greater atrophy. White matter hyperintensities (WMHs) were segmented using BISON^60^ as part of the PELICAN pipeline. Total WMH volume was calculated by summing voxel-wise WMHs and expressed in cubic millimeters. Given the typical right-skewed distribution of WMH volumes, values were log-transformed prior to statistical analyses to normalize their distribution. Quality control was conducted through visual inspection of all registration and segmentation outputs to identify and exclude failed cases.

### 2.4 Statistical analysis

Analyses were conducted separately within each cohort to preserve cohort-specific characteristics and account for differences in available measures and cohort structure. All continuous variables were standardized (z-scored) to facilitate comparison of effect sizes across models. Cross-sectional associations between education (or CR composite score in CIMA-Q), and cognitive outcomes were examined using linear regression models. All models included age at baseline and sex as covariates, and cognitive status was additionally included in CIMA-Q analyses to account for clinical heterogeneity.

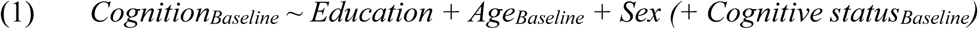

Associations between education (or CR composite score) and brain pathology markers were also examined:

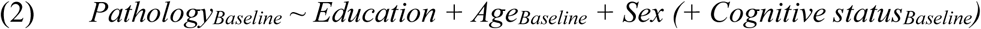

Fully adjusted multimodal models were subsequently constructed by including cerebrovascular, neurodegenerative, and AD biomarker measures alongside education (or CR composite score) and covariates, in order to determine whether associations between education (or CR composite score) and cognition remained statistically significant after accounting for combined pathology burden:

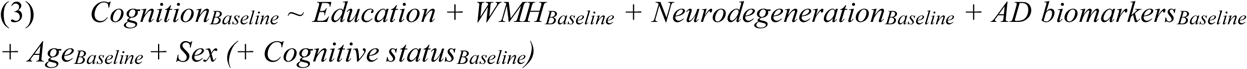

Longitudinal analyses were conducted using linear mixed-effects models to examine trajectories of cognitive change. Time was modeled as years from baseline assessment. Follow-up duration differed between cohorts. In PREVENT-AD, participants had a mean follow-up of 9.09 years, with a median of 9 visits (IQR: 6-13) and 4,648 total observations. In CIMA-Q, the mean follow-up was 2.44 years, with a median of 2 visits (IQR: 1-3.75) and 1,029 observations. Linear mixed-effects models with random intercepts and slopes were used to account for between-subject differences in baseline cognitive performance and within-subject variability in the trajectories of cognitive change. The interaction between education and time was used to determine whether education influenced the rate of cognitive decline. The model was specified as:

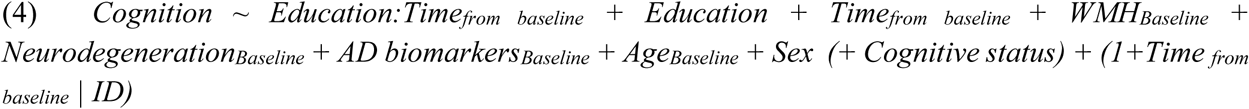

To account for multiple comparisons across models, false discovery rate (FDR) correction^61^ was applied using the Benjamini-Hochberg procedure, with statistical significance determined at an adjusted *p* value threshold of .05. All reported *p* values correspond to FDR-adjusted values. Additional analyses were conducted in the subset of CIMA-Q participants that were cognitively normal (CN) or SCD at baseline referred to as the CN group; to increase comparability across the two cohorts as PREVENT-AD classifies individuals with SCD as CN. This stratification allowed evaluation of whether associations differed across clinical stages and provided a subgroup more comparable to the cognitively unimpaired PREVENT-AD cohort. All analyses were performed in R (version 4.1.3), with *lme4* and *lmerTest* packages used for mixed-effects modeling.

### 2.5 Residual-based CR model

To further evaluate CR, residual-based analysis was conducted in both cohorts. This approach estimates cognitive performance (assessed by MoCA) relative to that expected from an individual’s level of brain pathology and demographic characteristics, such that positive residuals reflect better-than-expected cognition given pathological burden. To achieve this, a linear regression model was used with cognition as the dependent variable. Predictors included standardized measures of cerebrovascular, neurodegenerative, and AD biomarker measures, along with age, sex, and in CIMA-Q, cognitive status:

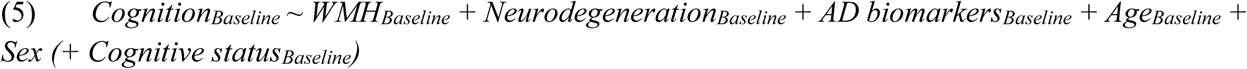

The residuals from this model were then extracted to derive an individual-level measure of CR, representing the discrepancy between observed and pathology-predicted cognitive performance. These residuals were subsequently standardized to generate a cognitive residual z-score. To examine whether education was associated with this residual-based CR measure, a second linear regression model was fitted with the cognitive residual score as the dependent variable and years of education as the independent variable, with age, sex, and cognitive status included as covariates where applicable. This analysis tested whether higher educational attainment was associated with greater residual-based CR beyond the effects of measured pathology and demographic covariates.

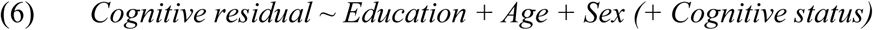

In addition, longitudinal analyses were used to examine whether education was associated with trajectories of residual-based CR change. Linear mixed-effects models were fitted with the cognitive residual as the outcome, including fixed effects of time, education, and their interaction, as well as random intercepts and slopes to account for within-subject variability:

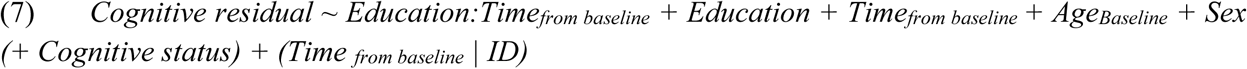

## 3. Results

### 3.1 Demographics and Clinical Data

A total of 432 and 422 participants were included from the PREVENT-AD and CIMA-Q, respectively. Participants in PREVENT-AD were significantly younger than those in CIMA-Q (63.6 ± 5.1 vs 73.95 ± 5.8 years, *p* < .001), while years of education were similar across cohorts. The proportion of females was also similar between PREVENT-AD and the full CIMA-Q sample. Baseline cognitive performance differed between cohorts, with higher MoCA scores observed in PREVENT-AD compared with CIMA-Q (28.0 ± 1.5 vs 25.7 ± 3.4, *p* < .001). Domain-specific cognitive measures also differed across cohorts, consistent with differences in clinical composition. In CIMA-Q, participants spanned multiple diagnostic groups, including cognitively normal individuals (CN: n = 68), SCD (n = 181), early and late MCI (n = 137), and mild AD (n = 36), reflecting a heterogeneous clinical sample. A CR composite score was available only in CIMA-Q (mean 16.4 ± 4.2, out of 25), reflecting multidimensional cognitive enrichment. Plasma biomarker levels differed across cohorts: plasma p-tau231 levels were higher in CIMA-Q than in PREVENT-AD (6.7 ± 3.2 vs 5.2 ± 2.9, *p* = .002), whereas plasma p-tau181 levels did not differ significantly between cohorts. Comparisons between PREVENT-AD and the CIMA-Q CN subgroup are reported in Table 1.

**Table 1.**
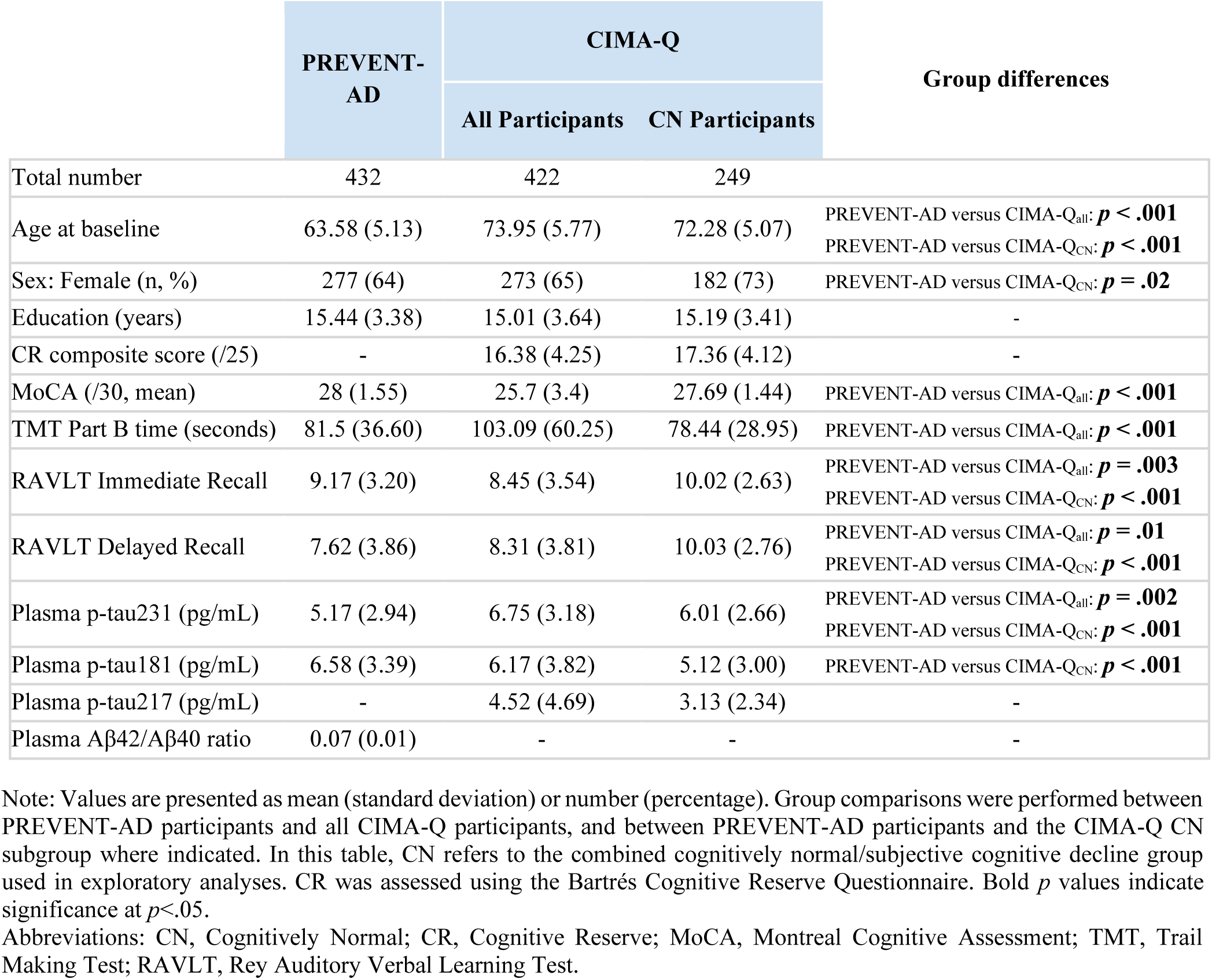
Cross-cohort comparison of demographic, cognitive, and biomarker measures.

|  | PREVENT-AD | CIMA-Q |  | Group differences |
| --- | --- | --- | --- | --- |
|  |  | All Participants | CN Participants |  |
| Total number | 432 | 422 | 249 |  |
| Age at baseline | 63.58 (5.13) | 73.95 (5.77) | 72.28 (5.07) | PREVENT-AD versus CIMA-Q<sub>all</sub>: ***p* < .001**<br>PREVENT-AD versus CIMA-Q<sub>CN</sub>: ***p* < .001** |
| Sex: Female (n, %) | 277 (64) | 273 (65) | 182 (73) | PREVENT-AD versus CIMA-Q<sub>CN</sub>: ***p* = .02** |
| Education (years) | 15.44 (3.38) | 15.01 (3.64) | 15.19 (3.41) | - |
| CR composite score (/25) | - | 16.38 (4.25) | 17.36 (4.12) | - |
| MoCA (/30, mean) | 28 (1.55) | 25.7 (3.4) | 27.69 (1.44) | PREVENT-AD versus CIMA-Q<sub>all</sub>: ***p* < .001** |
| TMT Part B time (seconds) | 81.5 (36.60) | 103.09 (60.25) | 78.44 (28.95) | PREVENT-AD versus CIMA-Q<sub>all</sub>: ***p* < .001** |
| RAVLT Immediate Recall | 9.17 (3.20) | 8.45 (3.54) | 10.02 (2.63) | PREVENT-AD versus CIMA-Q<sub>all</sub>: ***p* = .003**<br>PREVENT-AD versus CIMA-Q<sub>CN</sub>: ***p* < .001** |
| RAVLT Delayed Recall | 7.62 (3.86) | 8.31 (3.81) | 10.03 (2.76) | PREVENT-AD versus CIMA-Q<sub>all</sub>: ***p* = .01**<br>PREVENT-AD versus CIMA-Q<sub>CN</sub>: ***p* < .001** |
| Plasma p-tau231 (pg/mL) | 5.17 (2.94) | 6.75 (3.18) | 6.01 (2.66) | PREVENT-AD versus CIMA-Q<sub>all</sub>: ***p* = .002**<br>PREVENT-AD versus CIMA-Q<sub>CN</sub>: ***p* < .001** |
| Plasma p-tau181 (pg/mL) | 6.58 (3.39) | 6.17 (3.82) | 5.12 (3.00) | PREVENT-AD versus CIMA-Q<sub>CN</sub>: ***p* < .001** |
| Plasma p-tau217 (pg/mL) | - | 4.52 (4.69) | 3.13 (2.34) | - |
| Plasma Aβ42/Aβ40 ratio | 0.07 (0.01) | - | - | - |

Note: Values are presented as mean (standard deviation) or number (percentage). Group comparisons were performed between PREVENT-AD participants and all CIMA-Q participants, and between PREVENT-AD participants and the CIMA-Q CN subgroup where indicated. In this table, CN refers to the combined cognitively normal/subjective cognitive decline group used in exploratory analyses. CR was assessed using the Bartrés Cognitive Reserve Questionnaire. Bold *p* values indicate significance at *p* < .05.
Abbreviations: CN, Cognitively Normal; CR, Cognitive Reserve; MoCA, Montreal Cognitive Assessment; TMT, Trail Making Test; RAVLT, Rey Auditory Verbal Learning Test.

### 3.2. Cross-sectional associations between CR proxies and cognition

Higher education (as the main proxy of CR) was associated with better global cognition (assessed by MoCA) in age- and sex-adjusted models (Model 1) in both cohorts (PREVENT-AD: β = 0.15, *p* = .005; CIMA-Q: β = 0.27, *p* < .001). Similarly, higher CR composite score was significantly associated with better global cognition in CIMA-Q (β = 0.21, *p* < .001). Consistent associations were observed across cognitive domains, including executive function (TMT Part B) and episodic memory (RAVLT immediate and delayed recall), with positive associations between education and cognitive performance (Table 2). For the CR composite score in CIMA-Q, significant associations were observed for global cognition, executive function, and immediate memory, whereas the association with delayed recall was not statistically significant. Effect sizes for education were generally larger in the full CIMA-Q sample than in PREVENT-AD across corresponding models. The results were similar in the analyses that were restricted to CN participants in CIMA-Q. Education was associated with executive function and episodic memory outcomes, including TMT Part B time (β = 0.31, *p* = .01), RAVLT immediate recall (β = 0.24, *p* = .03), and RAVLT delayed recall (β = 0.20, *p* = .04), while the association with MoCA did not reach statistical significance (β = 0.15, *p* = .06). In the full CIMA-Q sample, effect sizes for education were slightly larger and more consistent than those observed for the CR composite score, particularly for global cognition and memory outcomes. Exploratory analyses of CR composite score subdomains showed that language proficiency, cognitive enrichment, and social engagement had smaller effect sizes than the total CR composite score and were not significantly associated with cognitive outcomes in age- and sex-adjusted models (Table S1).

**Table 2.** Cross-sectional associations between education or CR composite score and cognitive performance in age- and sex-adjusted models (Model 1)

|  | PREVENT-AD |  |  | CIMA-Q |  |  |  |  |  |  |  |  |
| --- | --- | --- | --- | --- | --- | --- | --- | --- | --- | --- | --- | --- |
|  |  |  |  | All Participants |  |  |  |  |  | CN Participants |  |  |
|  | Education |  |  | Education |  |  | CR Composite Score |  |  | Education |  |  |
| Cognitive Tests | $\beta$ | tStat | *p* | $\beta$ | tStat | *p* | $\beta$ | tStat | *p* | $\beta$ | tStat | *p* |
| MoCA | 0.15 | 2.82 | **.005** | 0.29 | 3.75 | **<.001** | 0.21 | 4.62 | **<.001** | 0.15 | 1.89 | 0.06 |
| TMT Part B time | 0.15 | 2.64 | **.008** | 0.24 | 2.38 | **.02** | 0.21 | 3.66 | **<.001** | 0.31 | 2.74 | **0.01** |
| RAVLT Immediate Recall | 0.17 | 3.04 | **.003** | 0.26 | 2.58 | **.01** | 0.15 | 2.52 | **.01** | 0.24 | 2.25 | **0.03** |
| RAVLT Delayed Recall | 0.12 | 2.29 | **.02** | 0.19 | 2.04 | **.03** | 0.11 | 1.88 | .07 | 0.20 | 2.14 | **0.04** |

Note: CR was assessed using the Bartres Cognitive Reserve Questionnaire. CN refers to the combined CN/SCD group in exploratory stratified analyses. For consistency across models, the TMT variable was reverse-coded such that higher values reflect better cognitive performance. All *p* values shown are FDR-adjusted. Bold *p* values indicate significance at $p < .05$ . Abbreviations: $\beta$ , standardized beta estimate; tStat, t statistic; CR, Cognitive Reserve; CN, Cognitively Normal; MoCA, Montreal Cognitive Assessment; TMT, Trail Making Test; RAVLT, Rey Auditory Verbal Learning Test.

Overall, cross-sectional analyses demonstrated consistent positive associations between education or CR composite score and cognitive performance across cohorts and domains. In fully adjusted multimodal models including cerebrovascular, neurodegenerative, and plasma biomarkers as covariates (Model 3), education remained significantly associated with cognitive performance in both cohorts (PREVENT-AD: β range: 0.11-0.15, all *p* ≤ .03; CIMA-Q all participants: β range: 0.18-0.27, all *p* ≤ .04) (Table 3; Figure 1). In CIMA-Q, the CR composite score was also associated with global cognition, executive function, and immediate recall (β range: 0.14-0.19, all *p* ≤ .04), whereas the association with delayed recall did not reach statistical significance (β = 0.10, *p* = .09). When analyses were restricted to CN participants in CIMA-Q, the results were more comparable to those observed in PREVENT-AD. In this subgroup, education was significantly associated with TMT Part B time (β = 0.25, *p* = .002) and RAVLT immediate recall (β = 0.23, *p* = .02), while associations with MoCA (β = 0.14, *p* = .06) and RAVLT delayed recall (β = 0.19, *p* = .06) did not reach statistical significance (Table 3). Exploratory fully adjusted analyses showed that individual CR subdomains were not significantly associated with cognitive outcomes and had small effect sizes (Table S2).

**Figure 1.**
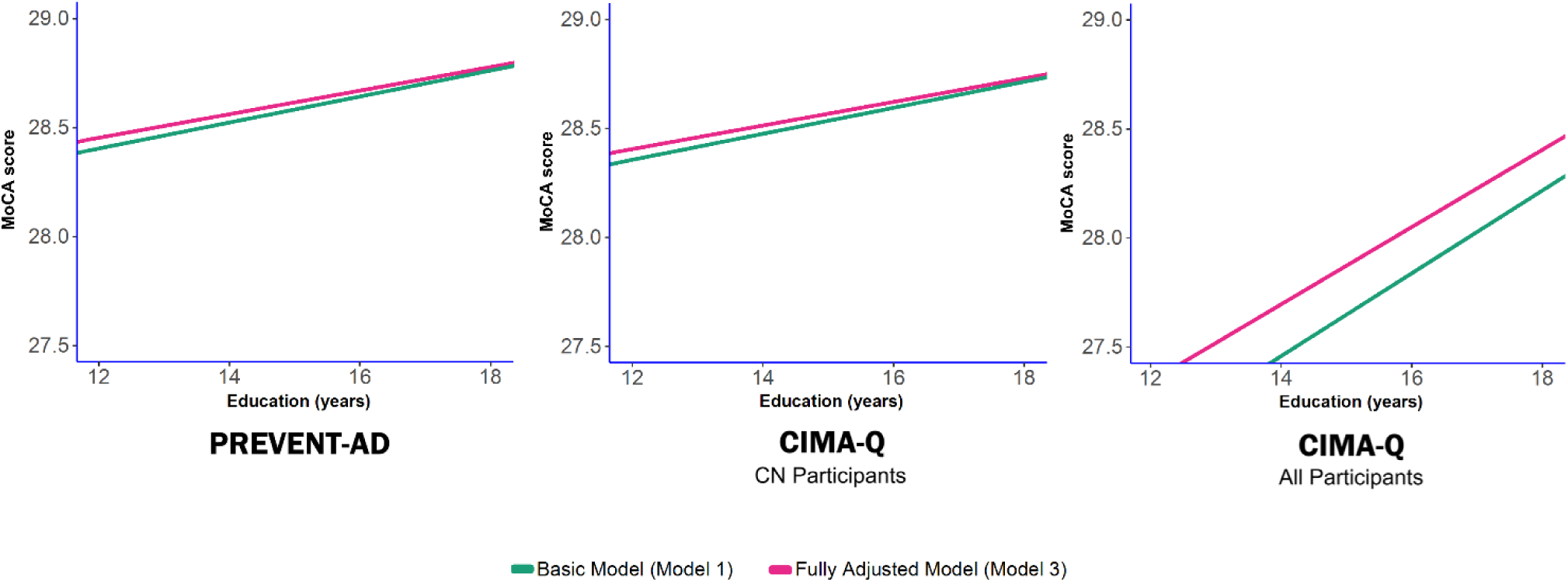
Association between education and cognitive performance across cohorts. MoCA scores are shown as a function of years of education in PREVENT-AD, CIMA-Q cognitively normal (CN) participants, and all CIMA-Q participants. Lines represent predicted values from the age- and sex-adjusted model (Model 1) and the fully adjusted multimodal model (Model 3). Model 3 additionally adjusted for cerebrovascular burden, neurodegeneration, and plasma Alzheimer’s disease biomarkers. Variables were standardized in the main analyses, but raw values are displayed here to improve interpretability.

**Table 3.**
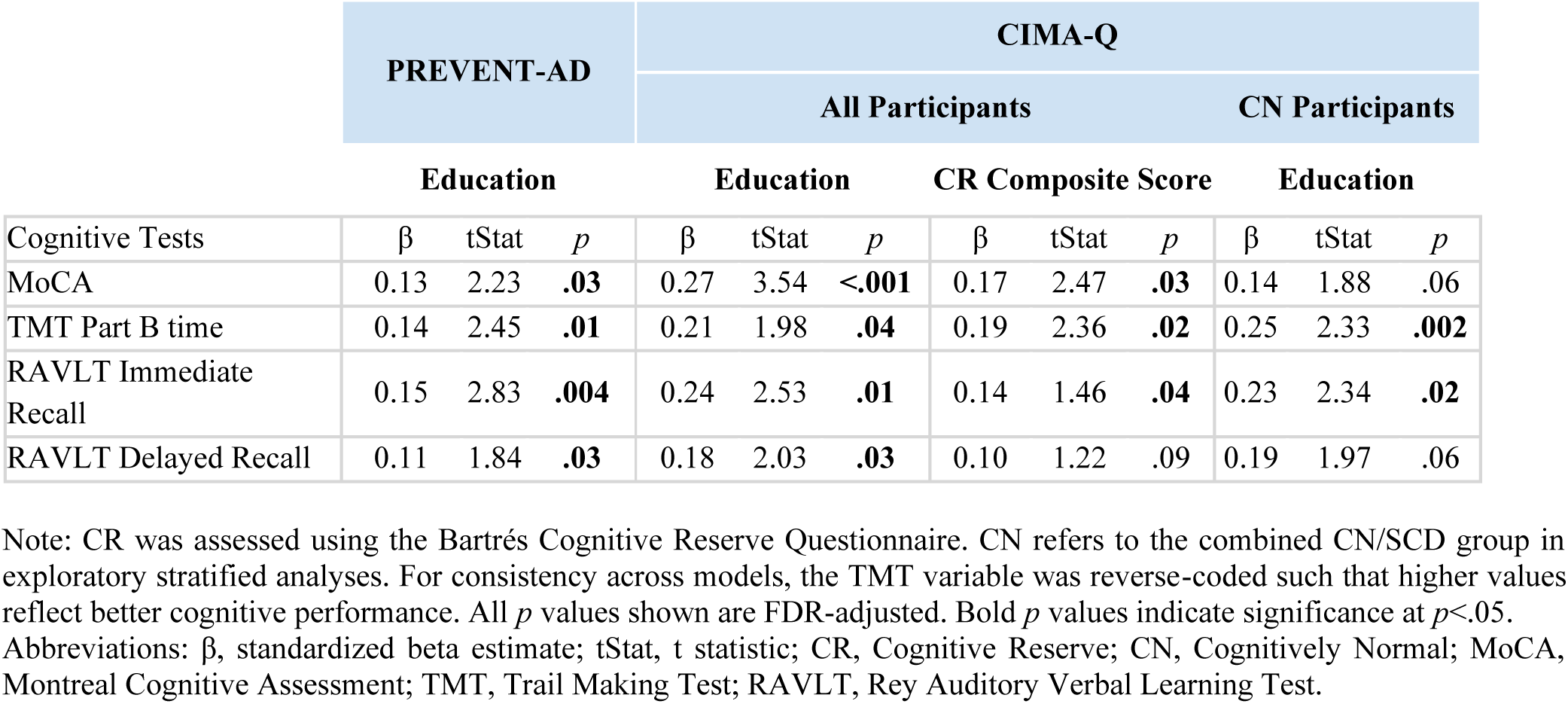
Cross-sectional associations between education or CR composite score and cognitive performance in fully adjusted multimodal models (Model 3)

The analyses were further repeated, adjusting for vascular, neurodegenerative, and plasma AD biomarkers individually (i.e., not included simultaneously), yielding similar results: higher education remained significantly associated with increased cognitive performance in both cohorts after adjusting for the other covariates in the model (PREVENT-AD: β range: 0.12-0.15, all *p* ≤ .03; CIMA-Q: β range: 0.23-0.27, all *p* ≤ .001). Similarly, higher CR composite was also significantly associated with increased cognitive performance in models including individual pathology markers (β range: 0.14-0.21, all *p* ≤ .02) (Table S3). CR subdomains again showed small and non-significant associations with cognitive outcomes (Table S4) in individually adjusted models.

### 3.3. Associations between CR proxies and multimodal brain measures

Cross-sectional analyses examining the association between education (or CR composite score in CIMA-Q cohort) and multimodal brain measures (Model 2) revealed no consistent evidence of an association between education and these measures across cohorts (Table S5). These measures reflect indices of cerebrovascular burden, neurodegeneration, and AD-related pathology derived from imaging and plasma biomarkers. This analysis was conducted to determine whether CR proxies were associated with lower measured brain pathology, which would be more consistent with brain maintenance or brain reserve, rather than CR mechanisms operating through better cognitive performance despite pathology. In PREVENT-AD, higher education was significantly associated with lower medial temporal lobe neurodegeneration (DBM; β = −0.09, *p* = .02), with a trend observed in association with the neurodegeneration composite measure (β = −0.05, *p* = .05). Education was not associated with cerebrovascular burden (WMH), ventricular atrophy, or plasma AD biomarkers (all *p* > .05). In CIMA-Q, education showed no significant associations with these brain measures (all *p* > .05), with the exception of a modest association with plasma p-tau231 (β = −0.19, *p* = .04). Similarly, the CR composite score was not significantly associated with any of these measures. These findings indicate that CR proxies showed limited associations with measured brain pathology. The analysis next examined whether these cross-sectional associations extended to longitudinal trajectories of cognitive change.

### 3.4 Longitudinal associations between education and cognition

Longitudinal follow-up times differed substantially between cohorts, with longer follow-up duration and more repeated assessments in PREVENT-AD compared to CIMA-Q (see Methods). In PREVENT-AD, fully adjusted linear mixed-effects models showed that education was positively associated with global cognitive performance (RBANS total: β = 0.27, *p* < .001) and with performance across multiple domains, including immediate memory, delayed memory, executive function, attention, language, and visuospatial ability (β range: 0.10-0.20, all *p* ≤ .04) (Model 4, Table 4). Education × time interactions were significant for executive function measures, including TMT total time (β = 0.09, *p* = .01) and TMT Part B time (β = 0.11, *p* = .01), but not for global cognition or other cognitive domains. In the TMT Part B trajectories, lower education was associated with a steeper decline over time, whereas higher education showed a more favorable trajectory, consistent with the significant education × time interaction.

**Table 4.**
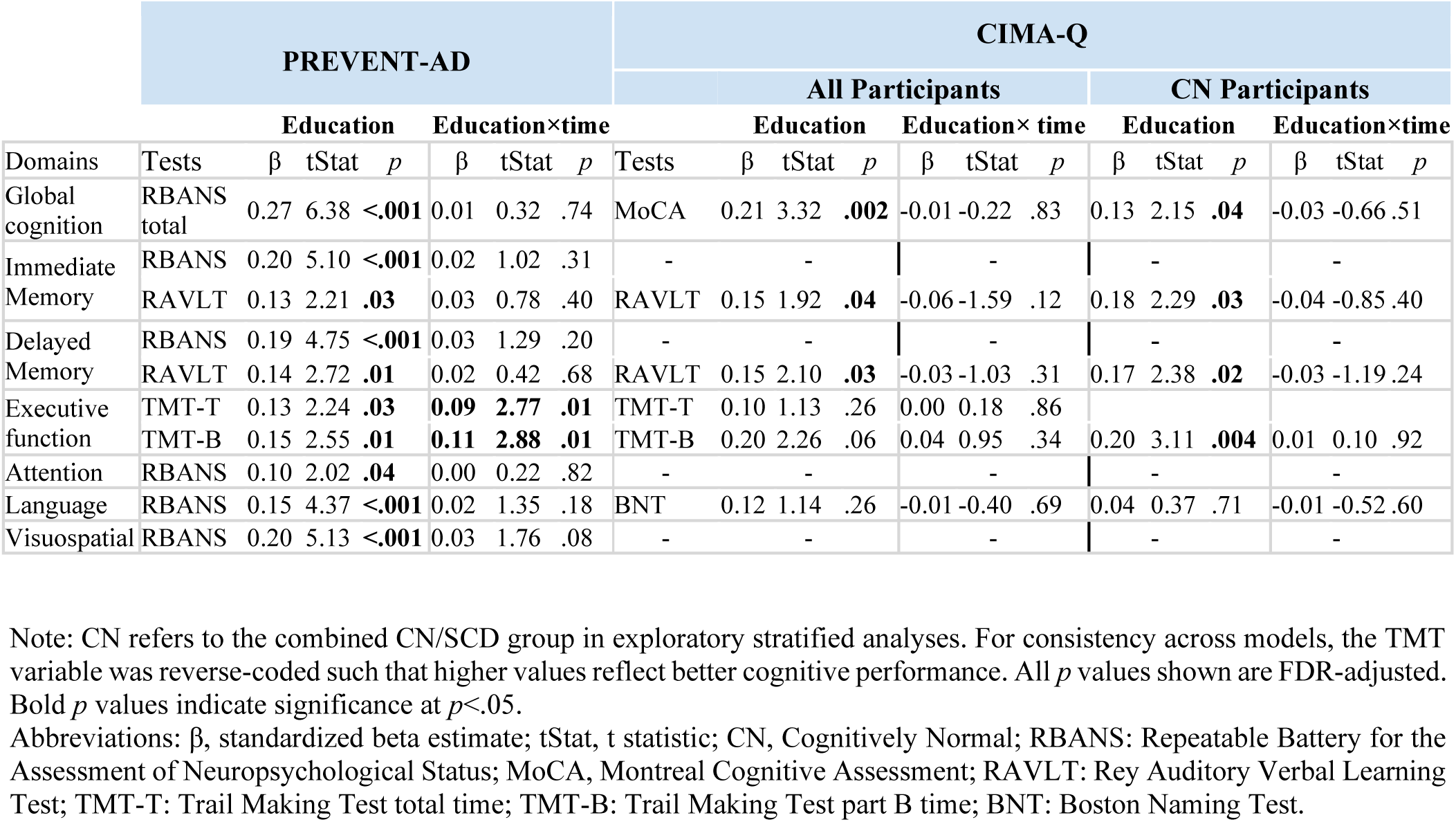
Longitudinal associations between education and cognitive performance and change (education × time) in fully adjusted multimodal models (Model 4)

In the full CIMA-Q sample, education was positively associated with global cognition (MoCA: β = 0.21, *p* = .002) and episodic memory measures, including RAVLT immediate recall (β = 0.15, p = .04) and RAVLT delayed recall (β = 0.15, *p* = .03). Associations with executive function and language measures were not statistically significant. Education × time interactions were not statistically significant for global cognition or any domain-specific outcome in the full CIMA-Q sample, however the shorter follow up might contribute to a limited range in change (Table 4). Predicted trajectories by education level are shown in Figure 2, and results from age- and sex-adjusted models without brain pathology measures are presented in Table S6. When CIMA-Q analyses were restricted to the CN group, education was associated with global cognition (MoCA: β = 0.13, *p* = .04), immediate recall (β = 0.18, *p* = .03), delayed recall (β = 0.17, *p* = .02), and TMT Part B time (β = 0.20, *p* = .004). However, unlike PREVENT-AD, education × time interactions were not significant in the CIMA-Q CN group.

**Figure 2.**
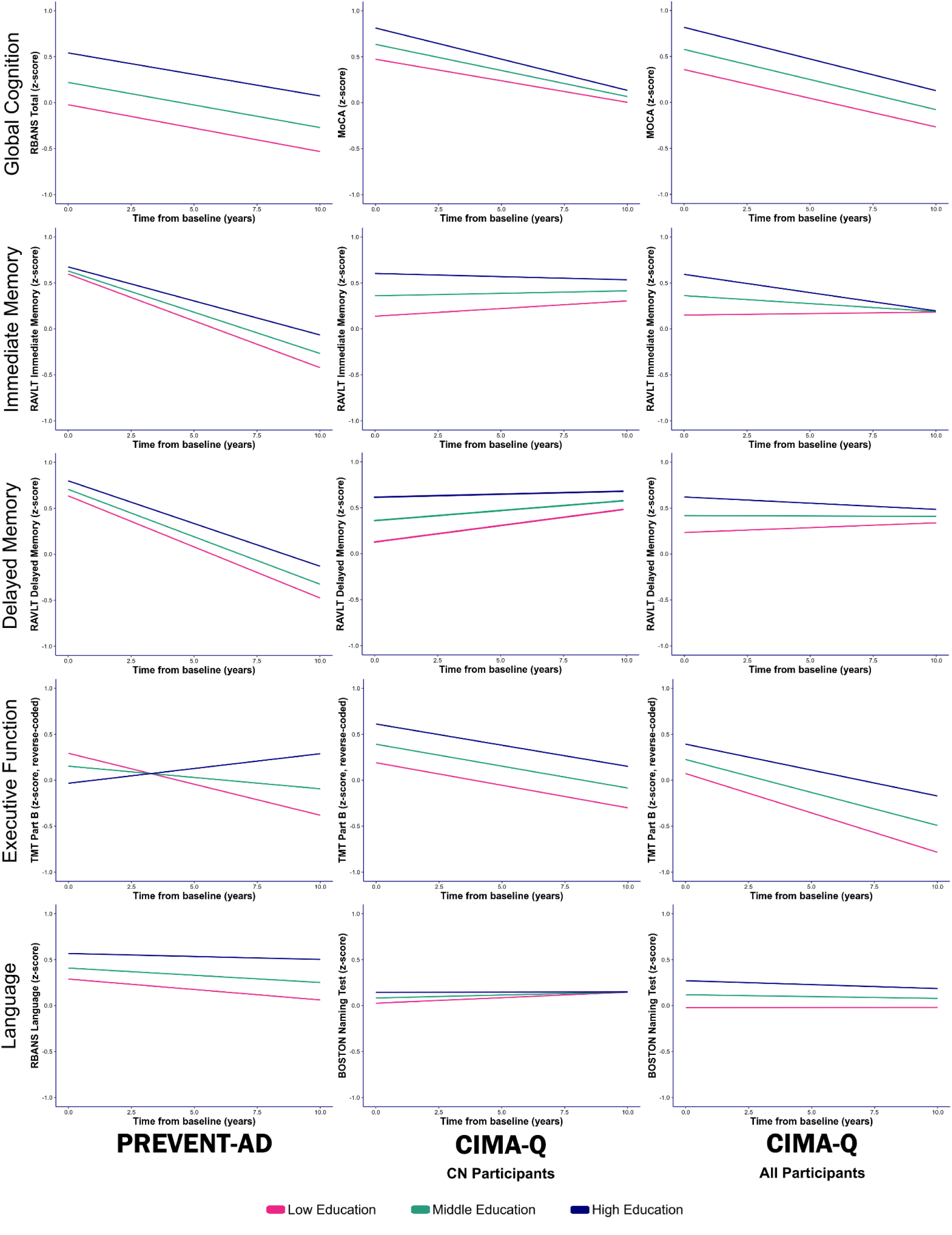
Longitudinal associations between education and cognitive performance across cohorts. Predicted cognitive trajectories are shown by education level in PREVENT-AD, CIMA-Q cognitively normal (CN) participants, and all CIMA-Q participants. Lines represent model-estimated values from fully adjusted linear mixed-effects models. Education was modeled as a continuous variable in all analyses but is displayed here in three levels for visualization purposes: low, middle, and high education. Cognitive outcomes are shown as z-scores; TMT Part B was reverse-coded such that higher values indicate better performance. Time is shown as years from baseline.

### 3.5 Residual-based CR

Residual-based CR analyses were conducted to assess cognitive performance relative to pathology-predicted performance. In cross-sectional analyses, higher education was significantly associated with greater residual-based CR in both cohorts (Model 6). In PREVENT-AD, education was positively associated with the cognitive residual score (β = 0.12, *p* = .04). In CIMA-Q, a larger association was observed (β = 0.45, *p* < .001). In contrast, the CR composite score was not significantly associated with the residual-based cognitive measure in CIMA-Q (β = 0.17, *p* = .18). Longitudinal residual-based analyses (Model 7) showed that education was significantly associated with higher residual-based cognitive performance in both cohorts (PREVENT-AD: β = 0.11, *p* < .001; CIMA-Q: β = 0.41, *p* < .001). Education × time interactions were not statistically significant in either cohort (both β ≈ 0.00, both *p* > .05), indicating no significant association between education and change in residual-based cognitive performance over time (Table S7).

## 4. Discussion

This study examined how CR proxies, including education, a multidimensional CR composite score, and a residual-based cognition measure, relate to cognitive performance and multimodal brain pathology across two cohorts representing complementary stages of the AD continuum. Across analytical approaches, higher education was the most consistent CR proxy associated with better cognitive performance after accounting for cerebrovascular, neurodegenerative, and AD-related pathology markers. In contrast, the multidimensional CR composite score showed less consistent associations, and exploratory analyses of CR subdomains showed small, non-significant effects. Longitudinally, higher education was more consistently associated with better sustained cognitive performance levels than changes in slopes of trajectories of cognitive change, although domain-specific effects were observed for executive function.

Our findings are consistent with extensive literature identifying education as a key proxy of CR^14,16–20,25,62^. Education remained associated with cognition after accounting for multimodal pathology, supporting a CR framework in which reserve-related factors contribute to cognitive performance beyond measurable structural brain changes^14,15,17,21^. This interpretation is further supported by residual-based analyses, in which education was associated with better-than-expected cognition given pathology burden^11,25,63^. At the same time, analyses of CR proxies and multimodal brain measures showed limited and inconsistent associations with measured pathology, suggesting that education-cognition associations are unlikely to be primarily explained by lower pathological burden^14,17,21,25^. This pattern is more consistent with CR or resilience mechanisms than with a purely brain maintenance explanation^6,25,63^.

Longitudinal analyses provided a more nuanced picture, suggesting that education-related CR was more consistently expressed as sustained higher cognitive performance than as slower cognitive decline^19,25^. Although early-stage groups in both cohorts showed education-related differences in executive-function performance, education-related differences in executive-function trajectories were observed only in PREVENT-AD. This finding may reflect the substantially longer follow-up duration and greater number of repeated assessments in PREVENT-AD, which likely improved sensitivity to detect subtle differences in within-person change^14,25^. The PREVENT-AD TMT Part B trajectories support this domain-specific interpretation. Lower education was associated with a steeper decline over time, whereas higher education showed a more favorable trajectory. This pattern suggests that education-related reserve may be particularly detectable in executive function, rather than reflecting a uniform effect across all cognitive domains^15,25,64^. This finding is biologically plausible because executive function depends on prefrontal and frontoparietal networks involved in cognitive control, flexibility, working memory, and processing speed^15,65–67^. These networks are highly plastic and responsive to lifelong cognitive stimulation, and prior studies have linked CR to executive functioning and more efficient recruitment of frontal networks^15,64,65^. PREVENT-AD participants were also younger and more likely to be in an active late-life stage, beyond chronological age alone, which may involve greater ongoing occupational, social, or cognitively demanding activities. Together, these factors may increase reliance on executive processes and make education-related differences in executive-function trajectories more detectable^68,69^. In contrast, memory trajectories showed less consistent education-related modification^14,16,19,25,70,71^. Episodic memory relies heavily on medial temporal lobe structures^12,26,72,73^, which are the earliest regions impacted by AD pathology^14,74^; once these regions are affected, compensatory capacity may be more limited, reducing the observable influence of CR^14,16,21,25^ .

Disease stage and availability of follow-up assessments likely contributed to the observed patterns. PREVENT-AD includes younger cognitively unimpaired individuals at elevated risk for AD^30,31^, whereas CIMA-Q includes a more heterogeneous population spanning normal cognition, SCD, MCI, and mild AD^37^. The comparison between PREVENT-AD and the CIMA-Q CN group is therefore particularly informative because both groups represent earlier clinical stages. Education was associated with executive function in both early-stage groups, supporting the robustness of this domain-specific association. However, the absence of significant education × time interactions in the CIMA-Q CN group may reflect the smaller sample size, shorter follow-up durations, and fewer repeated assessments, reducing our ability to detect subtle within-person changes.

Our findings also raise an important methodological point about CR measurement. Education showed the most consistent associations across cross-sectional, longitudinal, and residual-based analyses^19,21,22,25,63^, whereas the multidimensional CR composite score showed more variable associations and was not associated with residual-based CR. When education-related information was separated from the multidimensional questionnaire, the remaining CR subdomains, including language proficiency, cognitive enrichment, and social engagement, showed very small and non-significant associations with cognitive outcomes. This finding suggests that the non-education components of the multidimensional CR measure provided limited added explanatory value in the present analyses. Therefore, the evidence for CR in this study is driven primarily by education and residual-based cognition rather than by independent effects of questionnaire subdomains. This interpretation does not imply that multidimensional CR is unimportant, but suggests that its measurement is complex and requires further investigation. Education may act as a stable proxy because it captures early-life cognitive enrichment and is also linked to broader socioeconomic, cultural, and health-related exposures^11,17,19,20,25^. In contrast, questionnaire-based domains such as social engagement, leisure activity, music, or language may be more heterogeneous, context-dependent, and susceptible to measurement error or have restricted variability.

The integration of multimodal pathology represents a major strength of this study. Most prior research has examined CR in relation to single pathological markers^8,14,15,17,21,63^, whereas the current approach captures the combined influence of vascular, neurodegenerative, and AD-related processes. This study further advances prior work by combining cross-sectional models, residual-based analyses, and longitudinal models within a unified analytical framework, while leveraging two cohorts spanning preclinical and symptomatic stages of AD. Together, these complementary approaches strengthen the main conclusion that education is associated with better cognitive performance despite measured pathology, whereas evidence for education-related differences in cognitive decline trajectory slopes is limited and domain-specific.

Several methodological limitations should be acknowledged. First, the CR questionnaire was not available in PREVENT-AD, preventing direct comparability of multidimensional CR across cohorts. Second, education is an imperfect measure of CR and may be influenced by socioeconomic and cultural factors^11,19,20^. Third, the observational nature of the analyses, and the cross-sectional design of several key models, limit causal inference^14,70,75^. Fourth, although plasma biomarkers are increasingly validated, they may not fully capture the complexity of AD pathology^17,21,63^. Finally, differences in cognitive assessments across cohorts may contribute to variability in domain-specific findings^64,75,76^. Additional limitations include the shorter follow-up and fewer repeated assessments in CIMA-Q. Despite these limitations, the findings reinforce the central role of CR in explaining inter-individual variability in cognitive aging and highlight the need to distinguish between cognitive level and cognitive change. Clinically, education-related reserve may be most visible as better cognitive performance despite pathology, particularly in executive-function measures, rather than as uniformly slower decline. Future studies should examine whether executive-function trajectories are especially sensitive to reserve mechanisms, using harmonized cognitive measures, longer follow-up, and richer life-course CR indicators.

In summary, education, as the primary proxy of CR, was consistently associated with better cognitive performance and residual-based CR after accounting for multimodal brain pathology. Across early-stage cohorts, education was associated with executive-function performance, and in PREVENT-AD, higher education was also associated with a more favorable executive-function trajectory over time. However, CR-related associations were generally more evident for baseline cognitive levels than for longitudinal cognitive change. The multidimensional CR composite and its non-education subdomains showed limited added value beyond education in this dataset. Overall, these findings support a CR framework in which education is associated with better cognition despite measured pathology, while emphasizing that CR effects are domain-specific and that CR proxies differ in their explanatory value.

## AUTHOR CONTRIBUTIONS

Shima Raeesi, Amelie Metz, Katherine Chadwick, Yashar Zeighami, Cassandra Morrison, and Mahsa Dadar were involved with the conceptualization and design of the work. Shima Raeesi, Yashar Zeighami, and Mahsa Dadar completed the analysis. Cassandra Morrison, Mahsa Dadar, Amelie Metz, Katherine Chadwick, Yashar Zeighami, and Shima Raeesi were involved with data interpretation. Shima Raeesi wrote the manuscript. Cassandra Morrison, Amelie Metz, Katherine Chadwick, Yashar Zeighami, Mahsa Dadar, and Shima Raeesi revised and approved the submitted version.

## ACKNOWLEDGMENTS

Data used in the preparation of this article were obtained from the PRe-symptomatic EValuation of Experimental or Novel Treatments for Alzheimer’s Disease (PREVENT-AD) program data release 8.0. PREVENT-AD was launched in 2011 as a $13.5 million, 7-year public-private partnership using funds provided by McGill University, the Fonds de Recherche du Québec – Santé (FRQ-356162), an unrestricted research grant from Pfizer Canada, the J.L. Levesque Foundation, the Douglas Hospital Research Centre and Foundation, the Government of Canada, the Canada Fund for Innovation, the Canadian Institutes of Health Research (SV: 178385, JP: 153287, 178210, LC: 165921, TS: 175328,) the Alzheimer Society of Canada, the National Institutes of Health of the United States (NS: NIH AG068563), the Alzheimer Association (SB: AARG-NTF 926696) and Brain Canada Foundation. Parts of the data used in this article were obtained from the Consortium for the Early Identification of Alzheimer’s disease (CIMA-Q; https://www.cima-q.ca/en/home/). The CIMA-Q investigators contributed to the design, protocols and implementation of the study, as well as the collection of clinical, cognitive and neuroimaging data and biological samples. A complete list of the CIMA-Q investigators can be found at https://www.cima-q.ca/en/home/. The CIMA-Q is supported by the Fonds de recherche du Québec–Santé (FRQS) - Pfizer Innovation Program, FRQ cohort funds, the Quebec Network for Research on aging, the Fondation Courtois (NeuroMod project), the Consortium for the Neurodegeneration associated with Aging, and the Fondation Famille Lemaire. This study was supported by research funds from the Canadian Institutes of Health Research (CIHR). Dr. Dadar reports receiving research funding from the Fonds de Recherche du Québec-Santé (FRQS, https://doi.org/10.69777/330750), the Natural Sciences and Engineering Research Council of Canada (NSERC), and Brain Canada. Dr. Morrison reports receiving research funding from CIHR and NSERC. Shima Raeesi reports receiving funding from Brain Health (BH), Vascular Training (VAST) Platform and Fonds de recherche du Québec-Santé (FRQS). The authors acknowledge use of ChatGPT-5 for refinement of language and sentence structures to enhance the manuscript’s overall clarity and readability. The authors also acknowledge use of the Digital Research Alliance of Canada (https://www.alliancecan.ca/en) computing resources in the current work.

## CONFLICT OF INTEREST STATEMENT

The authors declare no conflicts of interest.

## CONSENT STATEMENT

Informed consent was obtained in writing from each participant or their designated study partner.

## DATA AVAILABILITY STATEMENT

Data used in preparation of this article were obtained from the PRe-symptomatic EValuation of Experimental or Novel Treatments for Alzheimer’s Disease (PREVENT-AD) program, data release 8.0 (https://www.centrestopad.com/).

The CIMA-Q data are available to the research community to address research questions related to dementia and aging, upon approval by the User Access Committee. Access is limited to CIMA-Q members. However, membership is open to all scientists interested in dementia research. Detailed information on the procedure for submitting a data access request can be found on the CIMA-Q website (https://www.cima-q.ca/en/home/).

## Supplementary materials

**Table S1.** Cross-sectional associations between CR subdomains and cognitive performance in age-and sex-adjusted models (Model 1)

| CIMA-Q |  |  |  |  |  |  |  |  |  |
| --- | --- | --- | --- | --- | --- | --- | --- | --- | --- |
|  | Language Proficiency |  |  | Cognitive Enrichment |  |  | Social Engagement |  |  |
| Cognitive Tests | $\beta$ | tStat | *p* | $\beta$ | tStat | *p* | $\beta$ | tStat | *p* |
| MoCA | 0.03 | 0.86 | .39 | 0.06 | 1.58 | .11 | 0.04 | 1.16 | .24 |
| TMT Part B time | 0.07 | 1.47 | .14 | 0.05 | 1.04 | .30 | 0.07 | 1.38 | .17 |
| RAVLT Immediate Recall | 0.01 | 0.16 | .87 | 0.02 | 0.45 | .65 | 0.01 | 0.19 | .84 |
| RAVLT Delayed Recall | 0.00 | 0.04 | .97 | 0.02 | 0.41 | .68 | 0.03 | 0.69 | .47 |

Note: CR subdomains were derived from items of the Bartrés Cognitive Reserve Questionnaire. Language proficiency reflects multilingual ability; cognitive enrichment reflects cognitively stimulating activities, including musical engagement, training courses, and intellectual activities; and social engagement reflects social participation and volunteer-related activities. For consistency across models, the TMT variable was reverse-coded such that higher values reflect better cognitive performance. Abbreviations: $\beta$ , standardized beta estimate; tStat, t statistic; CR, Cognitive Reserve; MoCA, Montreal Cognitive Assessment; TMT, Trail Making Test; RAVLT, Rey Auditory Verbal Learning Test.

**Table S2.** Cross-sectional associations between CR subdomains and cognitive performance in fully adjusted multimodal models (Model 3)

| CIMA-Q |  |  |  |  |  |  |  |  |  |
| --- | --- | --- | --- | --- | --- | --- | --- | --- | --- |
|  | Language Proficiency |  |  | Cognitive Enrichment |  |  | Social Engagement |  |  |
| Cognitive Tests | $\beta$ | tStat | *p* | $\beta$ | tStat | *p* | $\beta$ | tStat | *p* |
| MoCA | -0.04 | -0.70 | .49 | 0.06 | 0.07 | .30 | 0.00 | 0.08 | .94 |
| TMT Part B time | 0.08 | 1.23 | .22 | 0.13 | 1.83 | .07 | 0.00 | 0.02 | .98 |
| RAVLT Immediate Recall | 0.07 | 0.73 | .47 | 0.15 | 1.62 | .11 | -0.01 | -0.17 | .86 |
| RAVLT Delayed Recall | 0.05 | 0.59 | .58 | 0.11 | 1.27 | .21 | 0.07 | 0.85 | .39 |

Note: CR subdomains were derived from items of the Bartrés Cognitive Reserve Questionnaire. Language proficiency reflects multilingual ability; cognitive enrichment reflects cognitively stimulating activities, including musical engagement, training courses, and intellectual activities; and social engagement reflects social participation and volunteer-related activities. For consistency across models, the TMT variable was reverse-coded such that higher values reflect better cognitive performance. Abbreviations: $\beta$ , standardized beta estimate; tStat, t statistic; CR, Cognitive Reserve; MoCA, Montreal Cognitive Assessment; TMT, Trail Making Test; RAVLT, Rey Auditory Verbal Learning Test.

**Table S3:** Cross-sectional associations between education or CR composite score and cognitive performance (MoCA) in individually adjusted models.

|  | PREVENT-AD |  |  | CIMA-Q |  |  |  |  |  |
| --- | --- | --- | --- | --- | --- | --- | --- | --- | --- |
|  | Education |  |  | Education |  |  | CR Composite Score |  |  |
| Pathology Measures | $\beta$ | tStat | *p* | $\beta$ | tStat | *p* | $\beta$ | tStat | *p* |
| WMH | 0.13 | 2.35 | **.02** | 0.24 | 4.21 | **<.001** | 0.14 | 2.35 | **.02** |
| Medial temporal lobe DBM | 0.12 | 2.12 | **.03** | 0.24 | 4.85 | **<.001** | 0.15 | 2.47 | **.01** |
| Ventricular DBM (global atrophy marker) | 0.13 | 2.31 | **.02** | 0.23 | 4.76 | **<.001** | 0.14 | 2.32 | **.02** |
| Neurodegeneration composite (DBM) | 0.12 | 2.13 | **.03** | 0.24 | 5.86 | **<.001** | 0.15 | 2.47 | **.02** |
| Plasma Aβ42/Aβ40 ratio | 0.14 | 2.76 | **.01** | - | | | - | | |
| Plasma p-tau181 | 0.15 | 2.81 | **.01** | 0.26 | 4.02 | **<.001** | 0.15 | 2.30 | **.02** |
| Plasma p-tau217 | - |  |  | 0.27 | 5.48 | **<.001** | 0.21 | 4.10 | **<.001** |
| Plasma p-tau231 | 0.15 | 2.70 | **.007** | 0.26 | 3.93 | **<.001** | 0.16 | 2.36 | **.02** |

Note: CR was assessed using the Bartrés Cognitive Reserve Questionnaire. Medial temporal lobe DBM represents a composite of hippocampal, parahippocampal, and entorhinal deformation-based morphometry (DBM) measures. Ventricular DBM represents a composite of DBM measures from lateral, third, and fourth ventricles. Neurodegeneration composite shows combined medial temporal and ventricular DBM (neurodegeneration index). For consistency across models, medial temporal lobe DBM and plasma Aβ42/Aβ40 were reverse-coded such that higher values reflected greater pathology. All *p* values shown are FDR-adjusted. Bold *p* values indicate significance at *p*<.05.
Abbreviations: WMH, white matter hyperintensity; DBM, deformation-based morphometry.

**Table S4:**
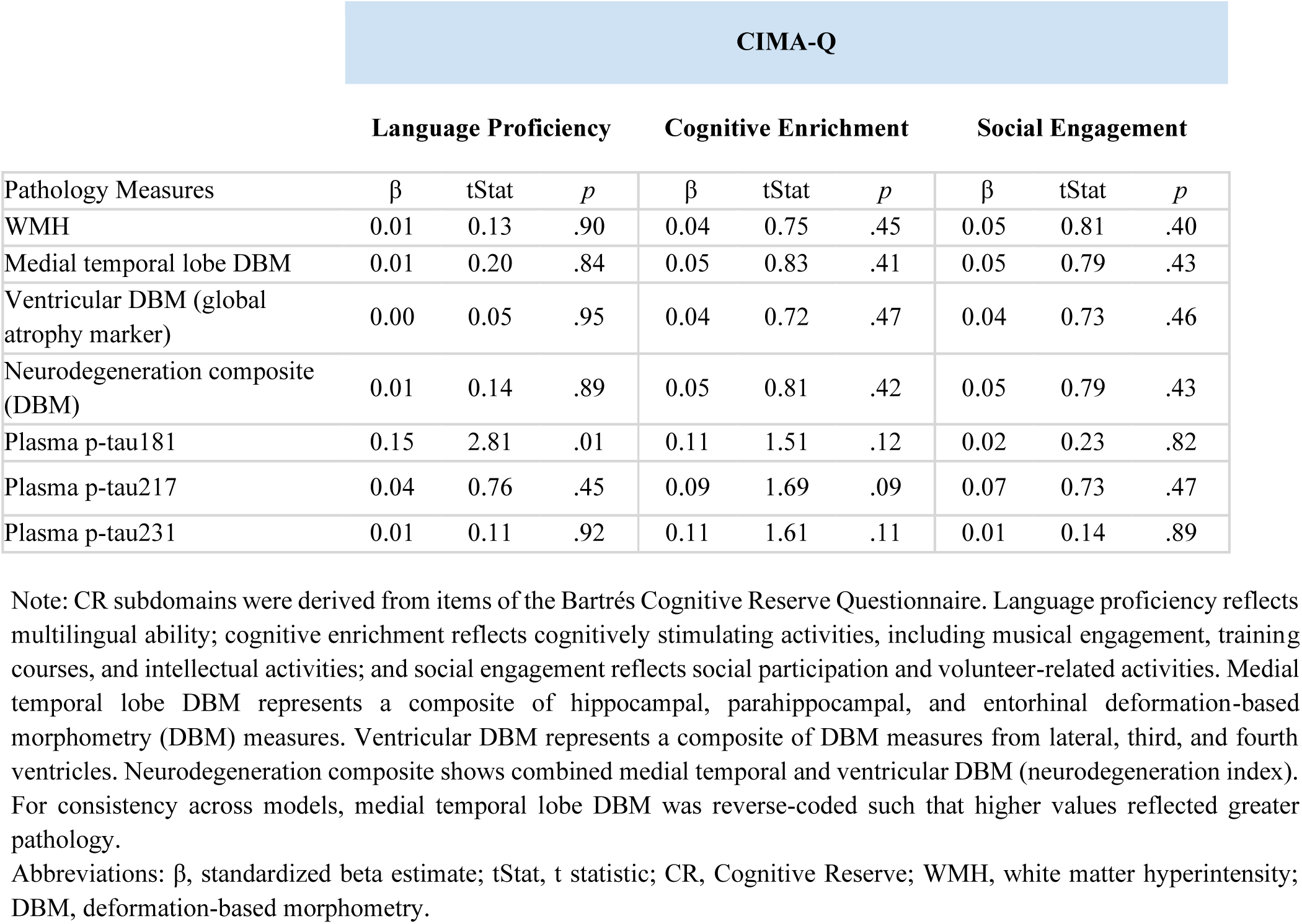
Cross-sectional associations between CR subdomains and cognitive performance (MoCA) in individually adjusted models.

**Table S5:**
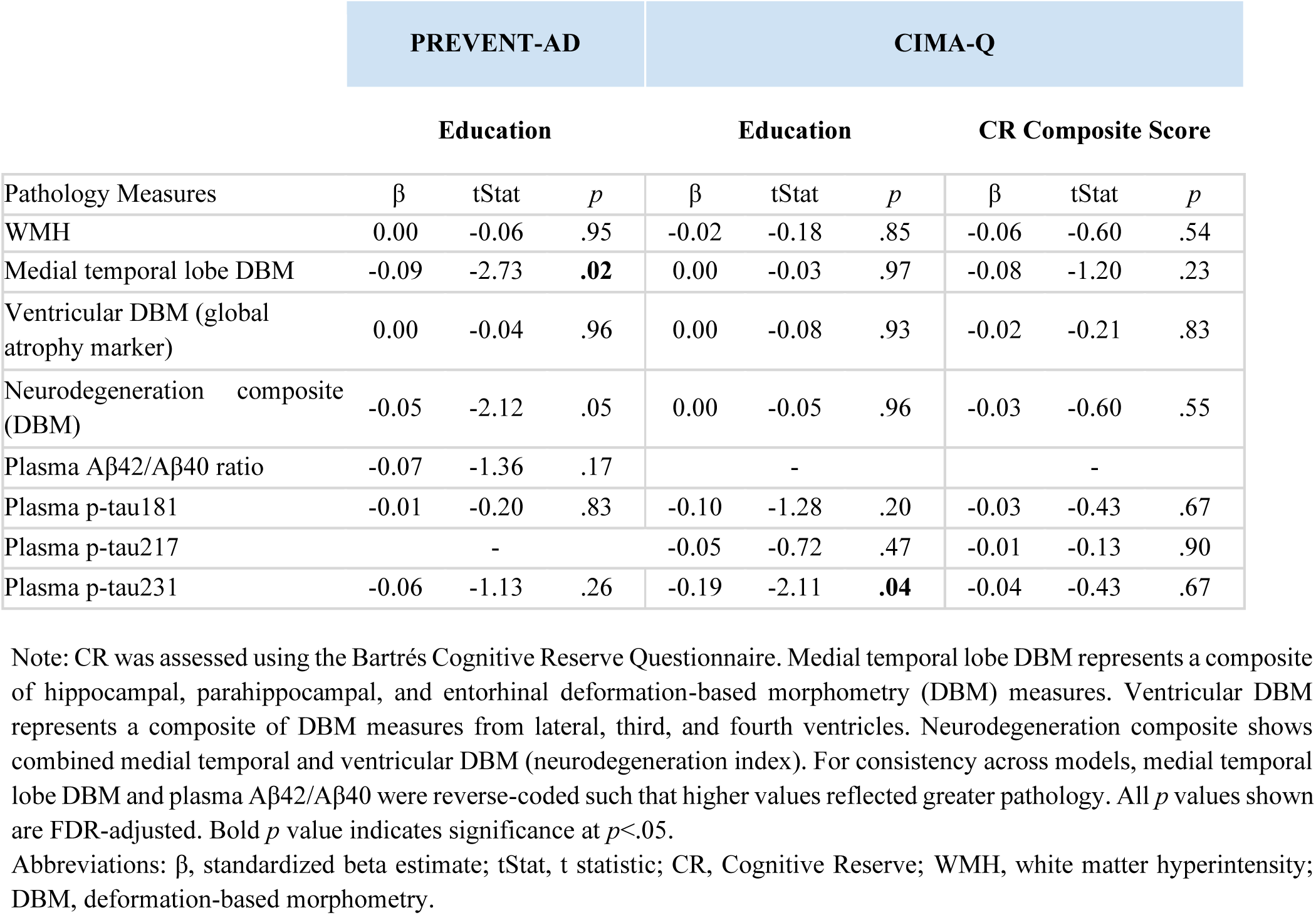
Cross-sectional associations between education or CR composite score and brain pathology measures (Model 2)

|  | PREVENT-AD |  |  | CIMA-Q |  |  |  |  |  |
| --- | --- | --- | --- | --- | --- | --- | --- | --- | --- |
|  | Education |  |  | Education |  |  | CR Composite Score |  |  |
| Pathology Measures | $\beta$ | tStat | *p* | $\beta$ | tStat | *p* | $\beta$ | tStat | *p* |
| WMH | 0.00 | -0.06 | .95 | -0.02 | -0.18 | .85 | -0.06 | -0.60 | .54 |
| Medial temporal lobe DBM | -0.09 | -2.73 | **.02** | 0.00 | -0.03 | .97 | -0.08 | -1.20 | .23 |
| Ventricular DBM (global atrophy marker) | 0.00 | -0.04 | .96 | 0.00 | -0.08 | .93 | -0.02 | -0.21 | .83 |
| Neurodegeneration composite (DBM) | -0.05 | -2.12 | .05 | 0.00 | -0.05 | .96 | -0.03 | -0.60 | .55 |
| Plasma Aβ42/Aβ40 ratio | -0.07 | -1.36 | .17 | - | | | - | | |
| Plasma p-tau181 | -0.01 | -0.20 | .83 | -0.10 | -1.28 | .20 | -0.03 | -0.43 | .67 |
| Plasma p-tau217 | - |  |  | -0.05 | -0.72 | .47 | -0.01 | -0.13 | .90 |
| Plasma p-tau231 | -0.06 | -1.13 | .26 | -0.19 | -2.11 | **.04** | -0.04 | -0.43 | .67 |

Note: CR was assessed using the Bartrés Cognitive Reserve Questionnaire. Medial temporal lobe DBM represents a composite of hippocampal, parahippocampal, and entorhinal deformation-based morphometry (DBM) measures. Ventricular DBM represents a composite of DBM measures from lateral, third, and fourth ventricles. Neurodegeneration composite shows combined medial temporal and ventricular DBM (neurodegeneration index). For consistency across models, medial temporal lobe DBM and plasma Aβ42/Aβ40 were reverse-coded such that higher values reflected greater pathology. All *p* values shown are FDR-adjusted. Bold *p* value indicates significance at *p*<.05.
Abbreviations: $\beta$ , standardized beta estimate; tStat, t statistic; CR, Cognitive Reserve; WMH, white matter hyperintensity; DBM, deformation-based morphometry.

**Table S6.** Longitudinal associations between education and cognitive performance and change (education × time) in age- and sex-adjusted models.

|  | PREVENT-AD |  |  |  |  |  |  | CIMA-Q |  |  |  |  |  |  |
| --- | --- | --- | --- | --- | --- | --- | --- | --- | --- | --- | --- | --- | --- | --- |
|  | Education |  |  |  | Education×time |  |  | Education |  |  |  | Education×time |  |  |
| Domains | Tests | β | tStat | *p* | β | tStat | *p* | Tests | β | tStat | *p* | β | tStat | *p* |
| Global cognition | RBANS total | **0.26** | **5.99** | **<.001** | 0.00 | 0.26 | .79 | MoCA | **0.20** | **3.19** | **.002** | -0.01 | -0.21 | .83 |
| Immediate Memory | RBANS | **0.18** | **4.38** | **<.001** | 0.02 | 1.00 | .32 | - | - | - | - | - | - | - |
|  | RAVLT | **0.12** | **2.08** | **.03** | 0.02 | 0.73 | .46 | RAVLT | 0.13 | 1.74 | .06 | -0.06 | -1.60 | .12 |
| Delayed Memory | RBANS | **0.17** | **3.99** | **<.001** | 0.03 | 1.28 | .20 | - | - | - | - | - | - | - |
|  | RAVLT | **0.14** | **2.64** | **.01** | 0.02 | 0.41 | .67 | RAVLT | **0.14** | **1.96** | **.05** | -0.03 | -1.04 | .31 |
| Executive function | TMT-T | **0.14** | **2.29** | **.02** | **0.09** | **2.77** | **.01** | TMT-T | 0.12 | 1.29 | .20 | 0.01 | 0.17 | .86 |
|  | TMT-B | **0.15** | **2.54** | **.01** | **0.11** | **2.86** | **.01** | TMT-B | 0.17 | 1.84 | .07 | 0.04 | 1.03 | .31 |
| Attention | RBANS | **0.11** | **2.33** | **.02** | 0.00 | 0.24 | .81 | - | - | - | - | - | - | - |
| Language | RBANS | **0.15** | **4.23** | **<.001** | 0.02 | 1.33 | .18 | BNT | 0.10 | 0.94 | .35 | -0.01 | -0.39 | .70 |
| Visuospatial | RBANS | **0.21** | **5.38** | **<.001** | 0.03 | 1.76 | .08 | - | - | - | - | - | - | - |

Note: For consistency across models, the TMT variable was reverse-coded such that higher values reflect better cognitive performance. All *p* values shown are FDR-adjusted. Bold *p* values indicate significance at *p*<.05.
Abbreviations: β, standardized beta estimate; tStat, t statistic; RBANS, Repeatable Battery for the Assessment of Neuropsychological Status; MoCA, Montreal Cognitive Assessment; RAVLT, Rey Auditory Verbal Learning Test; TMT-T: Trail Making Test total time; TMT-B: Trail Making Test part B time; BNT: Boston Naming Test.

**Table S7.** Cross-sectional and longitudinal associations between education and residual-based CR across cohorts.

|  |  | PREVENT-AD |  |  |  |  |  | CIMA-Q |  |  |  |  |  |
| --- | --- | --- | --- | --- | --- | --- | --- | --- | --- | --- | --- | --- | --- |
|  |  | Education |  |  | Education×time |  |  | Education |  |  | Education×time |  |  |
|  |  | β | tStat | p | β | tStat | p | β | tStat | p | β | tStat | p |
| Cross-sectional model |  | 0.12 | 2.06 | .04 | - |  |  | 0.45 | 3.73 | <.001 | - |  |  |
|  |  | Adjusted R-squared: 0.01 |  |  |  |  |  | Adjusted R-squared: 0.10 |  |  |  |  |  |
| Longitudinal model |  | 0.11 | 4.73 | <.001 | 0.00 | 0.5 | .62 | 0.41 | 7.33 | <.001 | 0.00 | 0.26 | 0.80 |
|  |  | Marginal R-squared: 0.05 |  |  |  |  |  | Marginal R-squared: 0.06 |  |  |  |  |  |

Note: Residual-based CR was defined as the standardized residual from models predicting MoCA from brain pathology and demographic covariates. Cognition was assessed using the Montreal Cognitive Assessment (MoCA) in cross-sectional models and in the CIMA-Q longitudinal analysis, whereas RBANS total score was used in the PREVENT-AD longitudinal analysis. All *p* values shown are FDR-adjusted. Bold *p* values indicate significance at *p*<.05.
Abbreviations: $\beta$ , standardized beta estimate; tStat, t statistic; MoCA, Montreal Cognitive Assessment; RBANS, Repeatable Battery for the Assessment of Neuropsychological Status.

## Notes

### Competing Interest Statement

The authors have declared no competing interest.

### Funding Statement

Dr. Dadar reports receiving research funding from the Fonds de Recherche du Quebec-Sante (FRQS, https://doi.org/10.69777/330750), the Natural Sciences and Engineering Research Council of Canada (NSERC), and Brain Canada. Dr. Morrison reports receiving research funding from CIHR and NSERC. Shima Raeesi reports receiving funding from Brain Health (BH), Vascular Training (VAST) Platform and Fonds de recherche du Quebec-Sante (FRQS).

### Author Declarations

The coordinating ethics committee of the Institut universitaire de geriatrie de Montreal gave ethical approval for the CIMA-Q study. Participants gave written informed consent before enrollment and for the different components of the project. PREVENT-AD data used in this study were obtained from an existing open-access dataset, the PRe-symptomatic EValuation of Experimental or Novel Treatments for Alzheimer's Disease program data release 8.0. The individual-level PREVENT-AD data had been de-identified prior to our use in this study. The PREVENT-AD dataset was prepared for data sharing through de-identification procedures, and access to sensitive participant-level data is provided through the registered PREVENT-AD repository under terms of use that prohibit any attempt to re-identify participants. The original PREVENT-AD study was conducted with informed consent and institutional ethics approval by the PREVENT-AD investigators.

